# Discovery of prognostic and treatment predictive biomarkers in recently diagnosed type 1 diabetes patients treated with anti TNFα

**DOI:** 10.1101/2023.05.29.23290668

**Authors:** S Kostense, Jaco M Klap, H Ashoor, R Yang, GJ Weverling, K Sweet, MR Rigby, JA Hedrick

**Affiliations:** Janssen R&D, Leiden, The Netherlands; Janssen R&D, Boston, MA, USA; Janssen R&D, Horsham, PA, USA

## Abstract

**Objective:** The T1GER study showed that treatment with the TNFα inhibitor golimumab in recently diagnosed type 1 diabetes patients showed better preservation of endogenous insulin production than placebo. However, considerable variation was observed among subjects. Therefore, a range of biomarkers were investigated for their potential to predict treatment response to golimumab.

**Research Design and Methods:** Baseline blood samples from 79 subjects were tested for autoantibodies, microRNA, metabolites, lipids, inflammatory proteins, and clinical chemistry. Univariate analysis was used to identify biomarkers that correlated with C-peptide change. Multivariate analysis was performed to establish a biomarker algorithm predicting the C-peptide response during the study.

**Results:** Multivariate analysis showed that baseline metabolites and miRNAs best predicted C-peptide responses both for placebo and treatment arms. Lipids, and inflammatory proteins were moderately predictive, whereas autoantibodies and clinical chemistry showed little predictive value.

An optimal model combining selected clinical variables and metabolites showed a correlation between predicted and observed C-peptide responses for the overall study up to 52 weeks, with an R^2^ of 0.85. An LOOCV model was developed as a surrogate validation test, resulting in an R^2^ of 0.69 overall, and an R^2^ of 0.76 specifically predicting C-peptide responses at week 38.

**Conclusions:** The exploratory analysis of the T1GER study resulted in a set of baseline biomarkers with promising performance in predicting future C-peptide responses during the study. If validated in independent cohorts, these prognostic and predictive biomarkers and algorithm carry significant translational impacts that can assist clinicians in making treatment decisions.

## Introduction

Type 1 diabetes is a chronic autoimmune disease of the pancreas that results in the destruction of insulin-producing beta cells (β-cells) of the islets of. Progressive destruction of β-cells eventually leads to metabolic imbalance and the inability to produce sufficient insulin for adequate glucose control without the use of exogenous insulin.

Although insulin therapy and blood glucose management provide substantial benefit to patients, this approach does not target the underlying destructive autoimmune processes that drive disease pathogenesis. Instead, immunomodulatory agents aimed to inhibit the beta cell specific autoimmunity, have shown to delay natural progression of the disease(1-3). The T1GER study showed C-peptide preservation in children and young adults with type 1 diabetes treated with the anti-TNFα antibody golimumab, compared to the placebo group (1). Almost half (41%) of the treatment group met one of the study’s definitions of responders, specifically higher or not more than 5% loss of C-peptide AUC at week 52 compared to baseline. Even so, in both the placebo and the treatment groups, large differences were observed in C-peptide responses during the study.

While the ability of golimumab treatment to delay disease progression is clear, the differential response to treatment warrants the investigation of predictive biomarkers that indicate which subjects will respond positively to treatment, and which subjects should avoid being exposed to potential side effects. Hereto baseline serum and plasma samples were collected from trial subjects and deployed multiple biomarker discovery technologies.

Type 1 diabetes is characterized by metabolic imbalance that is likely reflected in specific circulating metabolites (4-8). Lipidomics may harbor relevant biomarkers as well, especially in relation to TNFα which is known to affect lipid metabolism (9). Islet autoantibodies are a hallmark of the disease, and an autoantibody panel was included to investigate additional autoantibodies as biomarkers (10)). Inflammatory proteins were included due to the potential link with the disease specific islet inflammation (11), and the effect of the anti-TNFα antibody. In addition, miRNA’s were investigated as these have been implicated as beta cell death markers or general progression markers (12-15). Finally, clinical chemistry results were included as a negative control, as most of these measures are not expected to be linked to either the disease or treatment mechanism. This multi-omics approach was chosen for the discovery of both predictive biomarkers to determine which subjects respond to golimumab treatment, and prognostic biomarkers for the rate of disease progression in the placebo group.

## Materials & Methods

### Subject population and sample selection

Samples included in the current study were obtained from the T1GER study as described by Quattrin (1). The study was a Phase 2 multicenter, randomized-controlled placebo controlled trial. Individuals between 6-21 years old were enrolled within 100 days of their T1D diagnosis. Of the 84 participants, 56 were assigned to golimumab and 28 to placebo.

For the current evaluation, 1 placebo control and 4 treated subjects were excluded due to lack of follow up for C-peptide responses, resulting in 27 placebo and 52 treated subjects, 79 in total. The study design is shown in Figure 1A. Baseline samples were obtained during the screening period from 10 weeks before up until day 0 of the study start before treatment administration. Baseline serum or plasma samples were used for biomarker analysis as described below. Clinical end points, 4 hour C-peptide Area Under the Curve (AUC) upon a Mixed Meal Tolerance Test (MMTT), HbA1c, insulin use and Insulin Dose Adjusted HbA1c (IDAA1c) were determined at baseline, week 12, 26, 38, and 52.

**Figure 1.**
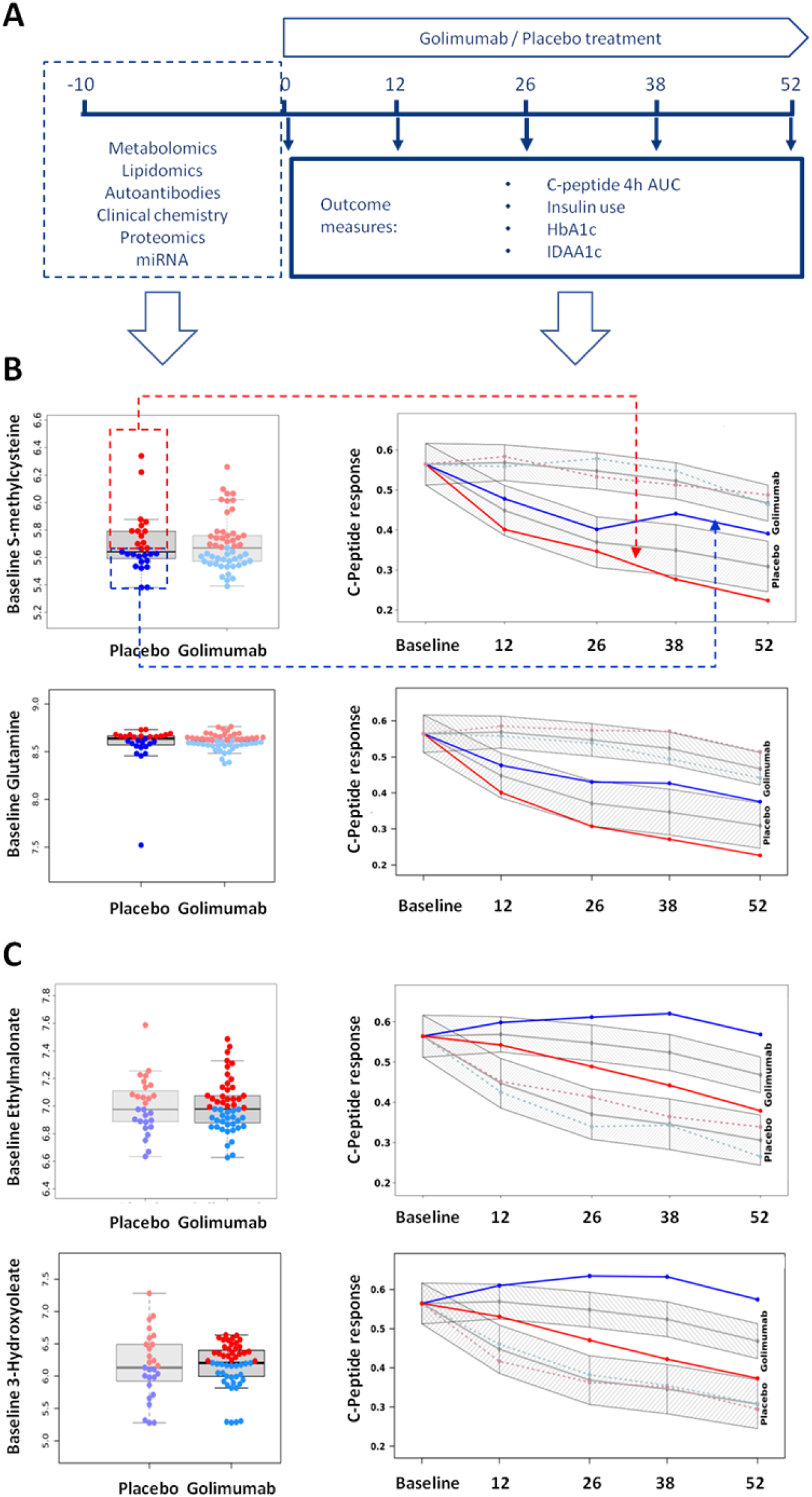
**A**: Study diagram. Recently diagnosed T1D patients were screened between -10 weeks and week 0, and serum and plasma samples were collected to perform multi omics biomarker analysis. Enrolled subjects were randomized to golimumab or placebo treatment at week 0 and were treated and followed up for the 52 weeks. At weeks indicated, blood samples were drawn to assess C-peptide responses upon MMTT.and HbA1c, insulin use was recorded and IDAA1c was determined as clinical outcome measures. Examples of baseline metabolites that function as **(B)** prognostic biomarkers for natural disease progression or **(C)** predictive biomarkers for treatment response. Dot plots in the left panels show the distribution of example metabolic biomarker levels at baseline for each subject in the study. Subjects are stratified in high (red), and low (blue) based on their biomarker level above or below the median. In the right panels the 4 hour C-peptide AUG (Ln(x+1)] is plotted over time, where the gray lines and areas indicate the overall C-peptide responses for placebo and treatment arms as reported for the T1GER study (Quattrin). Blue and red lines indicate the group averages of the stratified groups according to their baseline biomarker level (red=high; blue=low). Placebo arm subjects with low glutamine levels at baseline showed a slower decline in C-peptide response than placebo subjects with a high glutamine level at baseline. In the glolimumab treatment arm, subjects with a low baseline level of ethylmalonate showed a better C-peptide response upon treatment than subjects with a high baseline level of ethylmalonate.

### Metabolomics and lipidomics

For the global metabolomics analysis, performed by Metabolon (Morrisville, NC), plasma samples were extracted and split into equal parts for analysis on the three liquid chromatography tandem-mass spectrometry (LC-MS/MS) methods, and a polar LC method. For the CLP analysis, the samples underwent biphasic extraction and analyzed by LC-MS/MS in Multiple Reaction Monitoring (MRM) mode. Proprietary software was used to match ions to an in-house library of standards for metabolite identification and for metabolite quantitation by peak area integration.

### Proteomics

Protein analysis was performed on serum samples using the multiplex proximity extension assay (PEA) by Olink (Uppsala, Sweden). For this study 3 panels were selected: Inflammation, Cardiovascular III, and Cardiometabolic panels adding up to 276 proteins. For each panel, 1 μL sample was incubated with a mixture of 92 probe pairs, consisting of an antibody conjugated to a unique DNA oligonucleotide, in a 96-well plate. Pairs of antibodies bound to their corresponding antigens that form an amplicon by proximity extension of their DNA tails were quantified by high-throughput real-time PCR. Olink NPX Manager processing tool was applied to calculate Normalized Protein Expression (NPX) values, coefficient of variation and normalize data for subsequent statistical analysis.

### MicroRNA analysis

Serum samples were analyzed for the presence of microRNAs using the EdgeSeq technology by HTG Molecular (Tuscon AZ). The HTG EdgeSeq system combines HTG’s proprietary quantitative nuclease protection assay (qNPA) chemistry with a next-generation sequencing (NGS) platform to enable the semi-quantitative analysis of a panel of 2083 targeted miRNAs. Functional DNA nuclease protection probes (NPPs) flanked by universal wing sequences are hybridized to target RNAs. Universal DNA wingmen probes are hybridized to the wings to prevent S1 nuclease digestion. S1 nuclease is added to digest excess non-hybridized DNA probes and non-hybridized RNA, leaving only NPPs hybridized to RNA fully intact and able to be amplified / barcoded. Heat denaturation releases the protection probes from the DNA:RNA duplexes. The released DNA protection probes are ready for amplification / barcoding, quantification, and sequencing.

### Autoantibody analysis

Serum samples were analysed for autoantibodies using the KREX Immunome protein array by Sengenics (London, UK). This technology utilizes the biotin carboxyl carrier protein (BCCP) as a folding marker and solubility enhancer which results in expression of full-length, correctly folded and biotinylated proteins bound to the array slides. The immunome protein array consists of a panel of 1600 proteins to capture potential autoantibodies in the study samples.

### Clinical Chemistry

A total number of 52 clinical chemistry analyses were performed as per protocol in the T1GER study. Clinical chemistry analyses included general measurements such as liver enzymes, white blood cell counts, lipids, and antiviral antibodies.

### Statistical analysis

#### Univariate biomarker discovery

The estimators of the effects were derived by a repeated measurements model (week 0-week 52) that included baseline clinical variables, age, sex and BMI. A mixed-effect linear model was utilized to identify univariate biomarkers from our data to capture effect size at different visits while account for inter-visit correlations. The design of our model is:

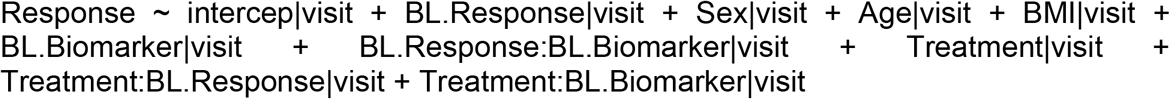

Where

BL.Response: Baseline response

BL.Biomarker: Biomarker level at the baseline timepoint

P-values were pooled across all visits to identify significant biomarkers. P-values corresponding to the BL.Biomarker|visit term represent prognostic biomarkers. P-values corresponding to the Treatment:BL.Biomarker|visit term represent predictive biomarkers for treatment effect.

### Multi-variate biomarker discovery

Group LASSO (least absolute shrinkage and selection operator) (16) was used coupled with recursive feature elimination (RFE) to identify the most predictive combination of biomarkers. A threshold value of unadjusted p<0.10 from the univariate analysis was used as an inclusion filter and standard L2 regularization searching for the optimal weight-decaying hyperparameter was applied to minimize cross-validation error (17). In the combined set of candidate predictors, LASSO (least absolute shrinkage and selection operator) downweighs the impact of predictors by using the λ_1_ penalty (17). A weight of 0 cancels out a candidate predictor from the model. Group LASSO was used to include or exclude a candidate predictor in a group-wise manner for the response at all the successive time points (16). The optimal penalization level was derived by minimizing the cross-validation error. The resulting set of predictors was consequently recycled though recursive feature elimination to yield only highly significant predictors.

## Results

### Univariate analysis of treatment predictive and prognostic biomarkers

In total, data was collected on 731 metabolites, 982 lipids, 2083 miRNA, 276 proteins, 1654 autoantibodies, 56 clinical chemistry assays on baseline serum or plasma samples, and 5 clinical variables from 79 subjects (Table I).

**Table I:** Result summary for univariate and multivariate analysis for each biomarker platform

| Biomarker platform | N detected biomarkers | N biomarkers in multivariate model after RFE | R<sup>2</sup> full multivariate model after RFE | R<sup>2</sup> LOOCV |
| --- | --- | --- | --- | --- |
| Metabolomics | 731 | 28 | 0.85 | 0.69 |
| MicroRNA | 2083 | 29 | 0.85 | 0.69 |
| Lipidomics | 982 | 20 | 0.69 | 0.48 |
| Proteomics | 276 | 17 | 0.67 | 0.44 |
| Autoantibodies | 1654 | 10 | 0.51 | 0.35 |
| Clinical chemistry | 52 | 8 | 0.50 | 0.38 |
| Clinical variables | 5 | 4 | 0.39 | 0.31 |
| Combined |  | 33 | 0.88 | 0.75 |

Pair-wise univariate analyses between measured baseline biomarkers and future clinical outcome measures were initially performed to identify prognostic and predictive biomarkers. Clinical outcomes included in these analyses were the MMTT induced C-peptide AUC, MMTT induced C-peptide max, HbA1C, Insulin use, and the Insulin Dose Adjusted for HbA1c (IDAA1C). C-peptide AUC was the major clinical outcome, including primary endpoint, reported for the T1GER study (1). Figure 1B and C show selected examples of metabolites that significantly predicted C-peptide responses during the study in either the placebo or treatment arm. Dot plots in the left panels show the biomarker levels at baseline for each subject per study arm. Each study arm was stratified into high and low based on the group median. The high (red) and low (blue) individuals are grouped, and their group average C-peptide response is plotted over time in the right panels. These show C-peptide responses (grey) as reported by Quattrin (1) using a [Ln(x+1)] transformation, and different subgroup trajectories with high (red) or low (blue) baseline biomarker levels. In the top panels of Figure 1B, placebo subjects with a high baseline level of S-methyl cysteine show a faster decline in C-peptide response than the low-level group. In the bottom panels, treated subjects with a low baseline level of 3-Hydroxyoleate, have a better C-peptide response upon treatment than the high-level subjects, whereas this pattern was absent in the placebo group.

### Multivariate analysis

Biomarkers predicting the C-peptide trajectory (mean C-peptide responses over time for a specific subgroup) for the pooled visits (unadjusted p value <0.05) or for at least one visit (unadjusted p value <0.01) from the univariate analyses were included for the multivariate analysis. Recursive feature elimination (RFE) to a performance of at least 95% of the optimal model was subsequently performed to identify the minimal biomarker set. The base model including only the clinical variables (Age, Sex, BMI, Treatment/Placebo, and Baseline C-peptide level) alone had limited predictive value (Figure 2A, left panel). The model based predicted C-peptide response on the y-axis only marginally correlates with the actual observed C-peptide responses on the x-axis with an R^2^ of 0.39. The model using 47 significant metabolites resulted in an R^2^ of 0.88 (Figure 2A 2^nd^ panel). Next we developed a model combining metabolites and clinical variables and consisting of 28 selected metabolites and 3 clinical variables that showed an R^2^=0.85 (Figure 2A, 3^rd^ panel). A list of the RFE selected metabolites and clinical variables is provided in the supplementary Table II.

**Figure 2:**
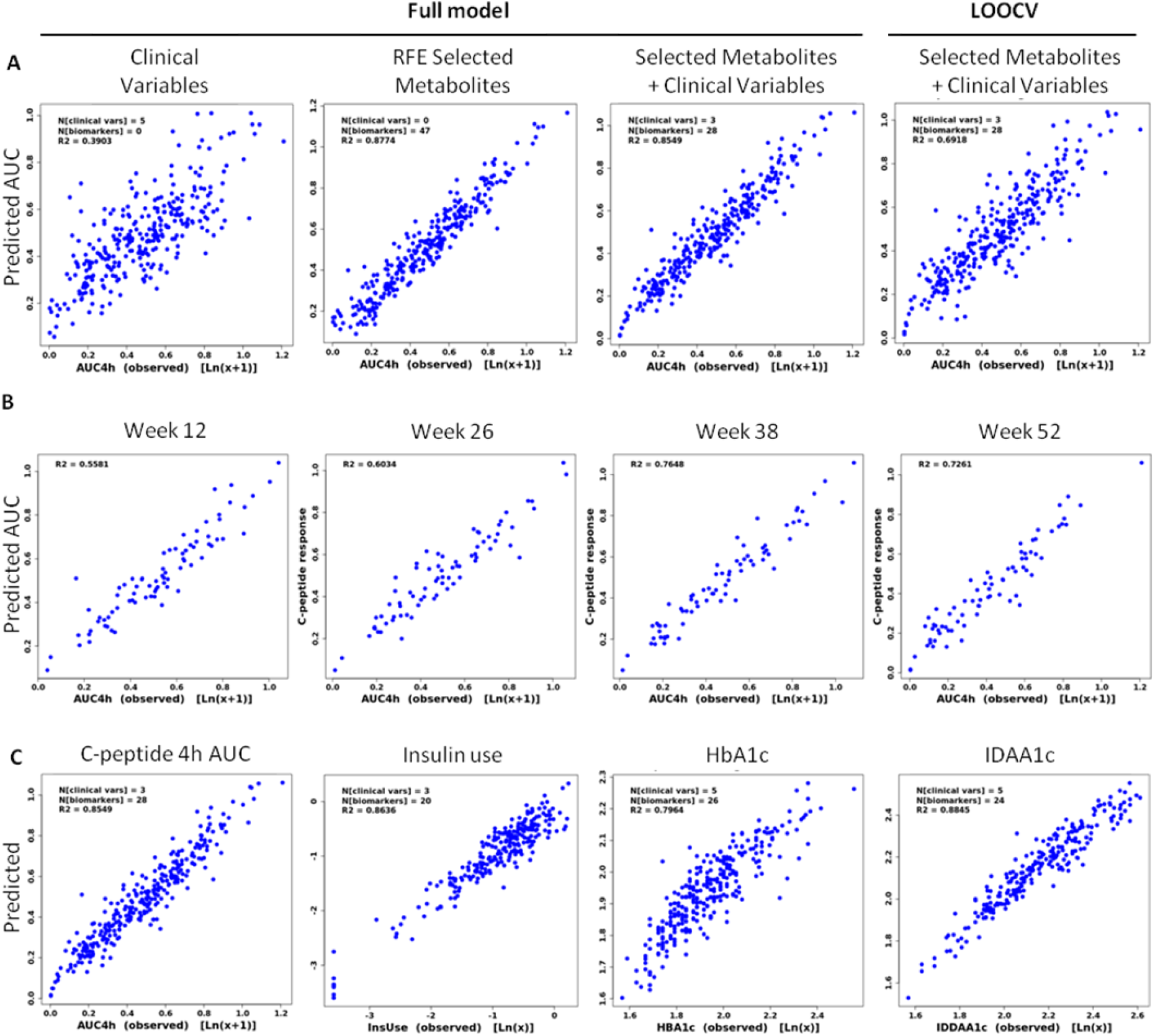
Multivariate model generation Model based predictions (Y-axis) are plotted against the C-peptide responses as observed in the trial (x-axis), in both placebo and treatment arms. The number of included clinical variables and metabolites at baseline included in the model is indicated in each panel in the top left corner. The R square indicates the agreement between predicted and observed data. **A:** Multivariate prediction models generated based on Clinical variables only. Metabolites only, a combination or Selected Metabolites and Clinical Variables, and the LOOCV model based on the Selected Clinical Variables and Metabolites. **B:** LOOCV multivariate model results for Selected Clinical Variables and Metabolites predicting c-peptide responses at week 12,26,38.and 52 respectively. **C:** Multivariate analyses were executed as described using selected clinical variables and metabolites predicting the four clinical outcome measures C-peptide, Insulin use, HbA1c, and IDAA1c.

For reference, the R^2^ for the complete set of biomarker platforms in combination with clinical variables are listed in Table I. Metabolomics and miRNA platforms yielded the best performance, followed by lipidomics and proteomics, and finally neglectable performance by autoantibodies and clinical chemistry.

### Model Validation using leave-one-out cross validation (LOOCV)

Given limited number of subjects per arm in the T1GER study, the Leave-One-Out CV was applied to assess model robustness. The LOOCV evaluation yielded to a respectable R^2^ of 0.69 (Figure 2A 4^th^ panel). The LOOCV results for the other biomarker platforms are listed in Table I, again showing similar performance robustness for the predictive effects of the multivariate analyses.

Next, we investigated for which timepoint in the study the model performed optimal. Figure 2B shows that the model performance at week 38 is the strongest R^2^ of 0.76. Similar results were obtained for the other biomarker platforms (data not shown).

While the C-peptide response was the key clinical end point for this analysis, models to predict the other clinical end points, such as HbA1c, insulin use and IDAA1c, were also developed. Figure 2C shows the full models after RFE for each of the end points. These results show that metabolites (and other biomarkers) can also be used to predict trajectories for other clinical outcomes.

A final analysis was done to combine all available biomarkers into a combined biomarker panel to predict the C-peptide response. This combination composed of 10 metabolites, 13 miRNAs, 3 lipids, 4 clinical chemistry assays, 2 proteins, 1 autoantibody and 3 clinical variables, and resulted in an R^2^ of 0.88 for the full model, and 0.75 for the LOOCV model.

This combination modestly improves the prediction model compared to the metabolomics model. Since feasibility of clinical application is greatly enhanced when using just one type of biomarker platform, further efforts were focused on metabolites.

### Applying the prediction model as a companion diagnostic tool

While the original report on the T1GER study the definition of a responder was set at less than 5% loss of C-peptide response at week 52, we aimed to visualize the difference between the choice for placebo versus golimumab treatment. Figure 3 shows the predicted C-peptide trajectories when chosen for placebo versus treatment for each individual subject in dotted lines. An overlay is plotted of the actual observed C-peptide responses for each subject, which is only for either the treatment or placebo option, depending on the subject randomization in the trial. The majority of predictions match nicely with the observed responses, reflecting the R^2^ 0.69 for the LOOCV model, with the exception noted in subject 10. Interestingly, the predictions sometimes indicate that treatment would result in worse C-peptide responses than placebo (subjects 20, 40); for many subjects there is no clear difference between treatment or placebo predicted (eg subjects 8, 23); for most subjects however, there is a clear treatment benefit predicted (eg subjects 31,33), in agreement with the overall results of the T1GER study(1).

**Figure 3.**
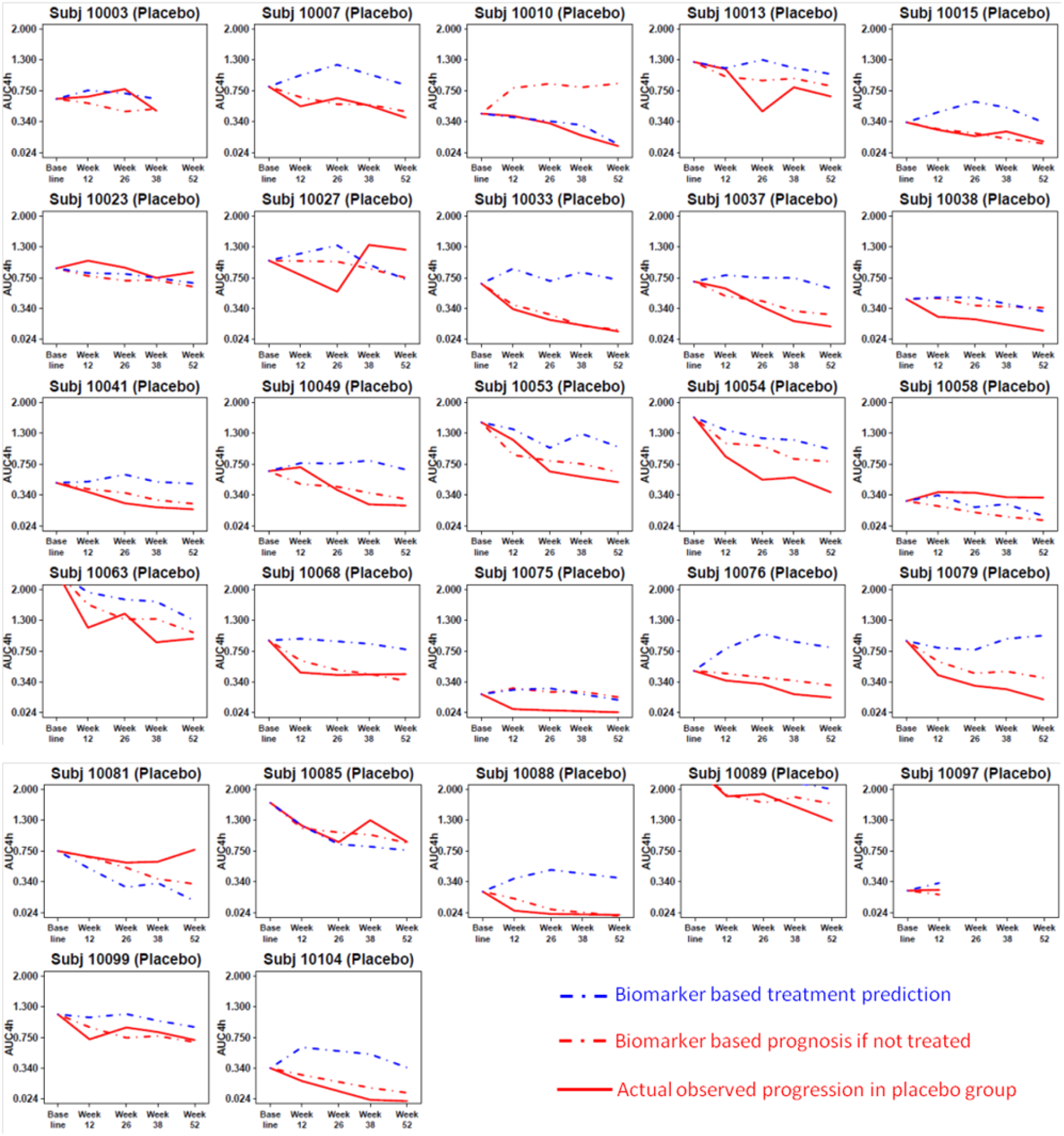
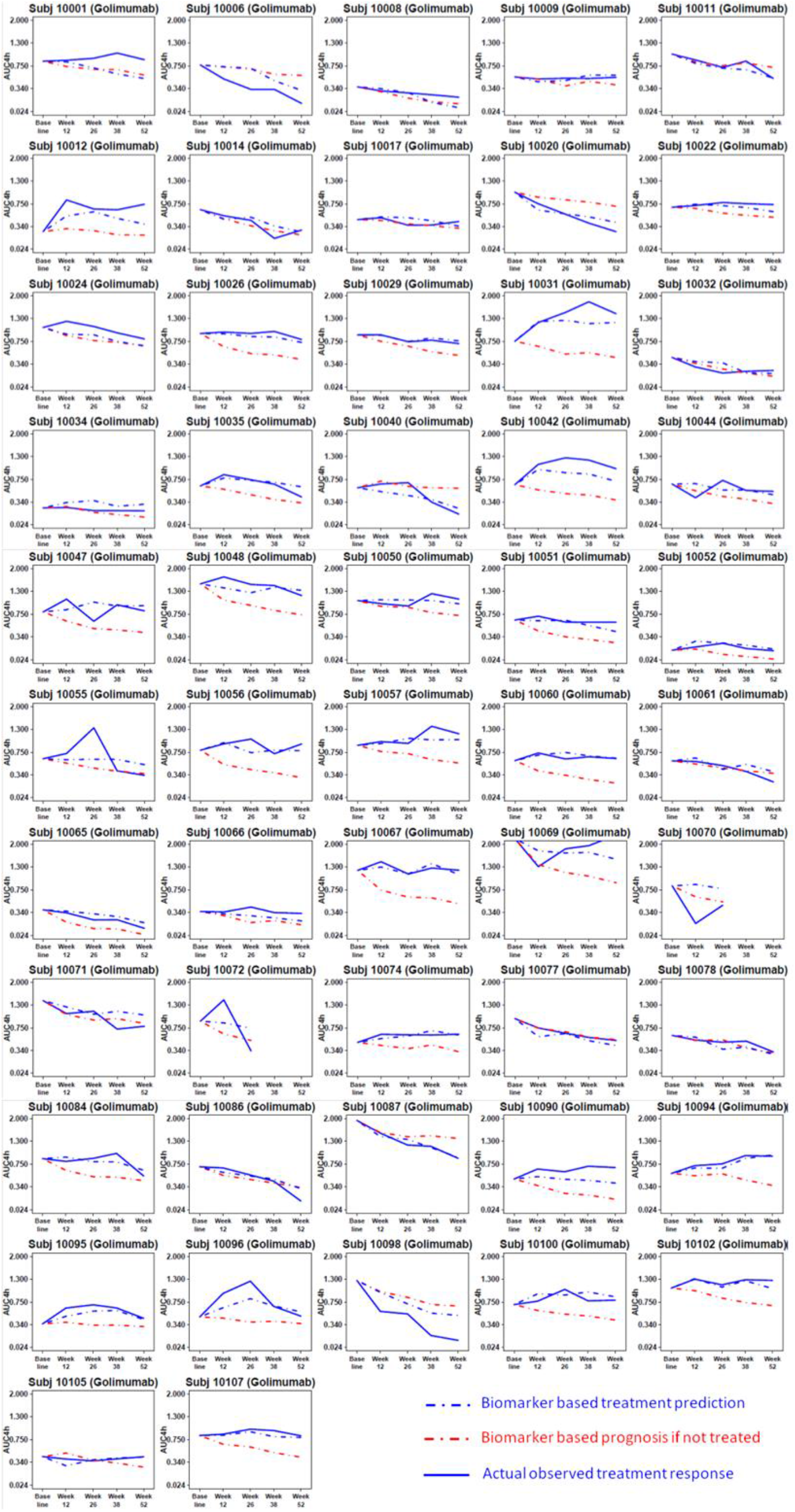
A: Individual subjects predicted vs observed C-peptide trajectories. Predicted C-peptide responses are plotted in dashed lines; observed in solid lines. Red: Placebo; Blue: Golimumab treatment. **A:** Placebo group; **B:** Treatment group

## Discussion & Conclusions

Decades of research in type 1 diabetes have resulted in a considerably improved understanding of the disease mechanism and has led to the emergence of immunomodulatory treatment to arrest beta cell destruction (1-3). However, similar to other autoimmune diseases, treatment is only successful in portion of treated subjects, leading to exposure of non-responders to the potential side effects of immunomodulatory therapy. Developing biomarkers to indicate which subjects would benefit from treatment remains a desirable goal in developing such therapies.

Here we report on the derivation of a prognostic and predictive biomarker panel, that was obtained in an agnostic approach using various omics technologies. Of the different biomarker types, metabolites and miRNA delivered the best signals for predicting treatment success in the treatment arm or disease progression in the placebo arm. Further development of this proposed biomarker panel will aid clinicians to make better informed treatment decisions, by which positive responders will be easier identified and selected for treatment, and fewer non responders will be exposed to potential side effects.

Given that the combination of different biomarker types only marginally improved the prediction compared to single biomarker platforms, a single biomarker platform may best more clinical implementable without compromising results.

There was no significant overlap in reported miRNA biomarkers in type 1 diabetes with our top performing miRNA’s. Previously reported miRNA markers for beta cell destruction MiR-375 (18; 19) and miR-204 (13) did not show up as significant biomarkers in our univariate analysis, nor did the miRNAs reported by Snowhite (15) that were associated with the C-peptide response at the 12 month MMTT.

We chose to focus on metabolomics-based model because, type 1 diabetes is an autoimmune disease with mainly metabolic consequences, and therefore the metabolomic platform would be a direct measure of the metabolic balance reflecting the disease state of an individual (4; 5; 20). Additionally, it could be argued that metabolites offer a better database to perform pathway analysis because metabolites can be directly linked to specific pathways, and miRNA often affect multiple pathways in epigenetics, making it more difficult to connect predictive miRNA with specific pathways. These are practical arguments and do not disqualify miRNAs as useful biomarkers.

TNFα blockers have been successfully used in various autoimmune related diseases (21). Despite increasing understanding of the molecular mechanism of TNFα and its blockers (22-24), it has proven difficult to develop treatment predictive biomarkers into a clinically applicable diagnostic test (25). Promising results have been reported using RNA sequencing to predict responses in RA (26; 27), cytokine profiling and transcriptomics in Spondyloarthiritis (28), and metabolic profiling in Ankylosing Spondylitis (29). The reported biomarkers in the listed studies do not clearly match with our findings, however, and these studies are difficult to compare due to differences in disease, biomarker technologies and study design. Overall, it appears that that TNFα affects multiple pathways in inflammation and lipid metabolism and more, which may also explain why different reports have found many different biomarker panels in TNFα treatment.

An interesting finding of our analyses is that the baseline biomarkers performed best when predicting the outcome at week 38. This may not be surprising as one can expect that treatment effect is still mounting at weeks 12 or 26 weeks, while after one year the effect may already be waning. A focus on week 38 may help to develop future models to be more accurate and meaningful.

While this study shows promising results, a caveat of this study is the size of the study, and the lack of validation of the biomarker panel and algorithm in an independent study. Biomarker discovery using multiple omics platforms on a limited study population does include the risk of overfitting and including non-relevant molecules by chance. Whereas we acknowledge this caveat, the difference in yield of predictive biomarkers in the different biomarker types -the ∼700 metabolites clearly outperforming the 1,600 autoantibodies-suggest that is not completely attributable to overfitting. As a surrogate validation exercise, an LOOCV model was developed on the same set of biomarkers included in the full model. This leave one out approach creates multiple models each used to predict the clinical end point for the subject who is left out. These models resulted in an R^2^ lower than the full model as can be expected but offer a preview of the performance that could be expected when the model is validated in an independent study. While the confirmation of the reported biomarker panel is dependent on the initiation of clinical trials investigating TNFα blockers in type 1 diabetes, the currently reported prognostic biomarkers for progression in the placebo group can be verified in natural progression studies or placebo arms of other clinical trials.

In the original report on the T1GER study(1), the definition of a responder was set at less than 5% loss of C-peptide (from baseline) at week 52. Whereas this is a simple and useful definition, it is not necessarily the best indication that a patient should be treated. A patient with low C-peptide response at baselinenot have much room for further decline and may show relatively stable C-peptide responses during the study, either with or without treatment. For instance, subjects 9, 17, 34, 66 in Figure 3 show stable C peptide responses during treatment and could be classified as responders who would benefit from treatment, but if no treatment is predicted to result in similar C-peptide levels, no treatment would actually be the better choice. Another example may be a treated patient who does show more than 5% loss of C-peptide, but without treatment would suffer a much worse decline of c-peptide response, and treatment may alleviate the otherwise fast progression (subjects 48, 53). In this study the biomarker panel was deployed to allow a comparison between treatment versus no treatment, instead of using a fixed cut point for the clinical end point.

The current study brings forward aspects that could improve clinical decision making for physicians deciding to treat type 1 diabetes patients with immunomodulatory therapy. We envision a future decision model in which baseline biomarkers can predict multiple clinical end points. For each individual patient, the physician could order a MMTT and a biomarker panel test. The algorithm can subsequently predict the trajectories for either treatment or no treatment. If the treatment prediction line is distinctively above the placebo predicted trajectory, the decision may favor treatment. If the difference is only marginal, no difference at all, or even worse for the treatment option, the physician should decide not to treat. With the current biomarker panel and algorithm developed in this study, this would already be possible. In this scenario, the decision is still subjective to the physicians opinion, but the algorithm could also include a calculation of the area under the curves of the prediction to obtain an objective difference between treatment or not over time, including costs of treatment. A further future development could include the biomarker-based prediction of insulin use, and HbA1c, adverse events linked to treatment, and type 1 diabetes related complications linked to HbA1c. All these predictions could feed a second algorithm that integrates the different predictions that can be translated in biological treatment effect in the form of beta cell function (C-peptide AUC), glucose control (IDAA1c), future risk for complications (HbA1c) etc, and offset these benefits with the cost for treatment and risk of side effects. While further validation is needed to enable this vision, this work sets the foundation for which this future may be built upon.

## Data Availability

All data produced in the present study are available upon reasonable request to the authors

## Authors contribution

S.K. designed the study, collected data and wrote the manuscript. J.M.K. designed the statistical analysis plan, performed statistical analysis and reviewed/edited the manuscript. H.A. performed statistical analysis and reviewed/edited the manuscript. R.Y. designed the statistical analysis and reviewed/edited the manuscript. G.J.W. designed the statistical analysis and reviewed the manuscript. K.S. designed the study, collected data and reviewed/edited the manuscript. M.R.R. contributed to the discussion and reviewed/edited the manuscript. J.H. contributed to discussion and reviewed edited the manuscript.

All authors were employees of Janssen R&D

Part of this study has been presented as an oral poster presentation at the Immunology of Diabetes Society (IDS) conference, November 2021

---

**Supplementary Table II:**
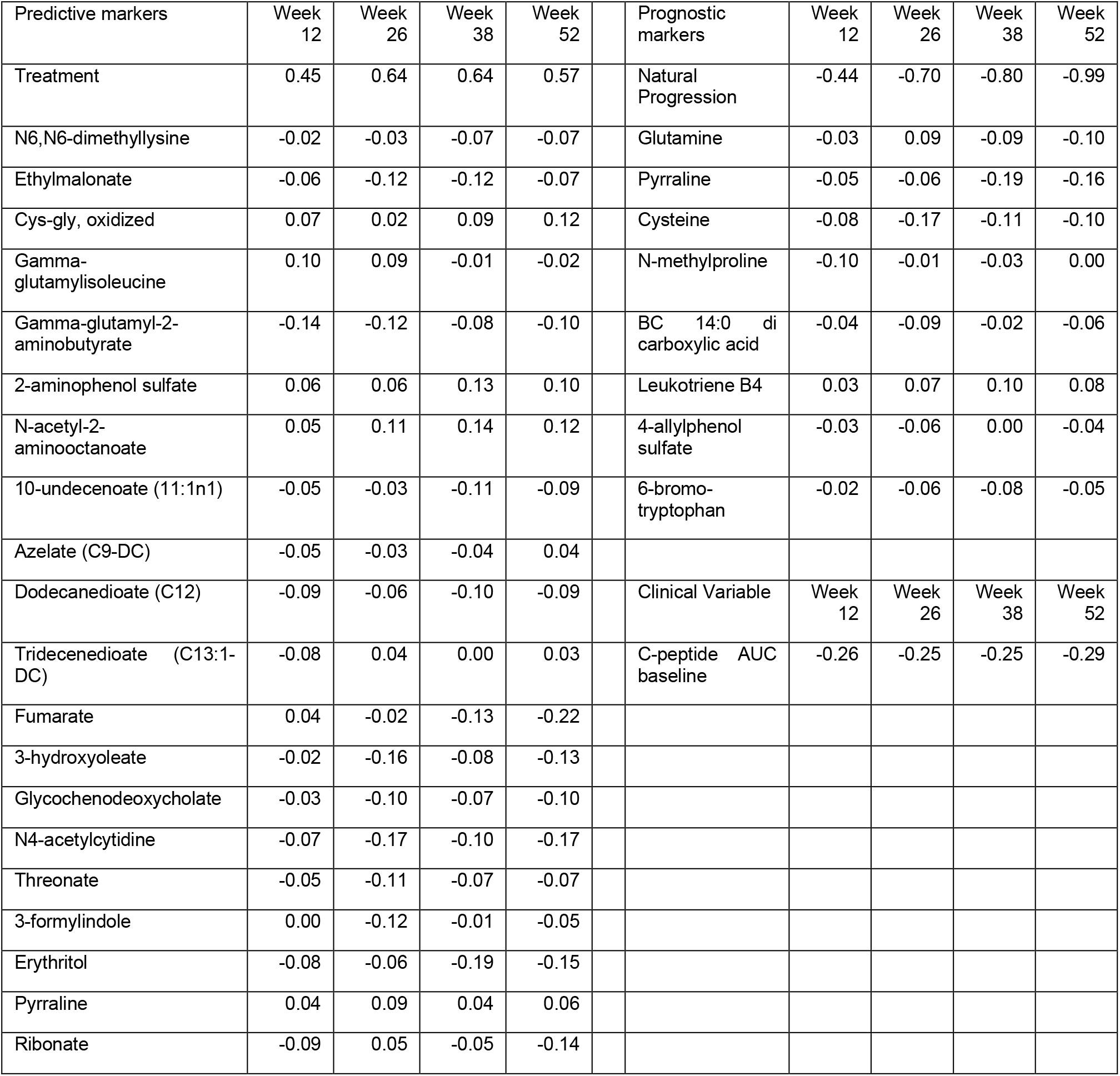
Multivariate model selected metabolites and clinical variables included in the full model

